# Predicting Harmful Alcohol Use Prevalence in Sub-Saharan Africa between 2015 and 2019: Evidence from Population-based HIV Impact Assessment

**DOI:** 10.1101/2024.03.24.24304804

**Authors:** Mtumbi Goma, Wingston Felix Ng’ambi, Cosmas Zyambo

**Author notes:** Corresponding author information: Mtumbi Goma, University of Zambia, Department of Public Health and Family Medicine, Lusaka, Zambia.

## Abstract

**Introduction:** Harmful alcohol use is associated with significant risks to public health outcomes worldwide. Although data on harmful alcohol use have been collected by population-based HIV Impact Assessment (PHIA), there is a dearth of analysis on the effect of HIV/ART status on harmful alcohol use in the SSA countries with PHIA surveys. This study uses data from the national representative PHIA to predict the harmful alcohol use prevalence.

**Methods:** A secondary analysis of the PHIA surveys: Namibia (n=27,382), Tanzania (n=1807), Zambia (n=2268), Zimbabwe (n=3418), Malawi (n=2098), Namibia (n=27,382), and Eswatini (n=2762). Using R version 4.2, the outcome variable and the descriptive variables were tested for association using chi square. Multivariable logistic regression analysis was used identify significant variables associated with harmful alcohol use. We employed to test and apply machine learning (ML) methods through Super Learner, Decision Tree, Random Forest (RF), Lasso Regression, Sample mean and Gradient boosting. Evaluation metrics methods specifically confusion matrix, accuracy, precision, recall, F1 score, and Area under the Receiver Operating Characteristics (AUROC) were used to evaluate the performance of predictive models. The cutoff point for statistically significant was P<0.05.

**Results:** Of the 12,460 persons, 15% used alcohol harmfully. Harmful alcohol use varied by countries and ranged from 8.7% in Malawi to 26.1% in Namibia (P<0.001). Females were less likely to use alcohol in a harmful way (AOR = 0.32, 95% CI: 0.29-0.35, P< 0.001). Compared to those HIV negative, persons that were with HIV-positive and on ART were less likely to use alcohol in a harmful way (AOR = 0.65, 95% CI: 0.57-0.73, P<0.001) however persons that were HIV-positive and not on ART were more likely to use alcohol in a harmful way (OR = 1.49, 95% CI: 1.32-1.69, P<0.001). Being married or formally married was protective to harmful use of alcohol. The best performing models were Lasso or Super Learner or Random Forest were the best performing models while gradient boosting models or sample mean did not perform well.

**Conclusion:** The findings highlight concerning variations in harmful alcohol use prevalence across surveyed countries, with Namibia reporting the highest rate. Males, older individuals, those HIV positive and not yet on ART, and unmarried persons demonstrated a higher likelihood of engaging in harmful alcohol use. These findings collectively contribute to a comprehensive understanding of the multiple factors influencing harmful alcohol use within the surveyed populations, the importance of targeted interventions at country and individual levels.

## INTRODUCTION

Over the centuries, alcohol has been used all over the world as an intoxicating drug that can lead to dependence. The consumption of alcohol is broadly embraced in social settings, often linked to feelings of relaxation and enjoyment. Despite some individuals being able to consume alcohol without adverse consequences, an increasing number of people encounter detrimental physical, social, and psychological effects associated with alcohol (1).

The prevalence of alcohol use and the problems associated with it vary greatly around the world, but the resulting effects on mortality and health are still present in many countries. On a global scale, harmful alcohol consumption is consistently ranked among the top five risk factors for illness, disability, and death (2).

Around 33.3 million deaths globally are attributed to harmful alcohol use each year, accounting for roughly 50.9 percent of all fatalities. Additionally, alcohol use is connected to 5 % of the world’s disease burden. More than 200 health conditions have been found to be causally linked to alcohol consumption in recent times. This includes new research showing a connection between harmful alcohol use and the incidence and clinical outcomes of infectious diseases like tuberculosis, HIV/AIDS, and pneumonia (2). In sub-Saharan Africa (SSA), where heavy alcohol consumption is common, harmful alcohol use is linked to disorders. According to estimates, the prevalence of alcohol use disorders is two to four times higher in people living with HIV (PLHIV) than in people without HIV infection. This increases the risk of HIV infection and transmission in a variety of contexts (3).

The harmful use of alcohol constitutes 5.1% of the worldwide burden of disease (National Collaborating Centre for Mental Health, 2011). Notably, harmful alcohol use emerges as the primary risk factor for premature mortality and disability in adults, contributing to 10% of all deaths globally. Additionally, disadvantaged and particularly vulnerable populations face elevated rates of alcohol-related deaths and hospitalizations (4).

Harmful alcohol use is associated with health status. In a longitudinal cohort study conducted in the Unites Sates of America, involving individuals with HIV who were engaged in care across seven clinics participating in the Centers for AIDS Research Network of Integrated Care Systems between January 2011 and June 2014 (n=5,046). The prevalence of heavy alcohol use was found to be 21% among women, 31% among men who have sex with women, and 37% among men who have sex with men. Among women, heavy alcohol use was associated with a subsequently decreased median self-reported health status score compared to those with no or moderate alcohol use (OR= 0.76; 95% CI: 0.58-0.99). This association was not explained by the presence of depressive symptoms. However, no significant association was observed between alcohol use level and subsequent self-reported health status among men who have sex with women. Among men who have sex with men, those reporting no alcohol use had a subsequently decreased median self-reported health status compared to those with moderate alcohol use (OR= 0.88; 95% CI: 0.80-0.97), (5). The findings from this longitudinal cohort study conducted in the United States highlight the significant association between harmful alcohol use and health status among individuals with HIV. The prevalence of heavy alcohol use varied across gender and sexual orientation groups, with notable impacts on self-reported health status.

In Ethiopia, the pooled prevalence of alcohol use among people PLWHIV was found to be high (6). In a cross-sectional community survey conducted on hazardous alcohol use and associated factors in a rural Ethiopian district, prevalence, and associated factors for hazardous drinking in rural Sodo district, southern Ethiopia were reported. The survey was part of a multi-center study, Programme for Improving Mental Health Care (PRIME), which is a consortium of research institutions and ministries of health of five low- and middle-income countries, namely Ethiopia, India, Nepal, South Africa and Uganda in partnership with UK institutions and World Health Organization (WHO). The cross-sectional community survey recruited 1500 adults, aged 18 and above, using multi-stage random sampling. Exploratory multivariable logistic regression was conducted to examine factors associated with hazardous alcohol use. Discussion of the paper revealed the need to integrate services for hazardous alcohol use such as brief intervention at different levels of primary care services in the district. The paper emphasized on the need to launch public health interventions to reduce hazardous alcohol use. Further, the findings highlighted the need for interventions to address alcohol use within HIV care and treatment programs in Sub-Saharan Africa (7).

Understanding the relationship between alcohol use and socioeconomic factors is important for developing effective public health interventions. Research by Kaushalendra Kumar highlights on the relationship between socio-demographic characteristics and harmful alcohol consumption in India. Kumar’s findings indicated that individuals aged over 50 were less likely to be heavy drinkers, emphasizing age as a negative correlation. Furthermore, being female was a significant protective factor against current drinking. Individuals having more than 12 years of schooling exhibited a reduced likelihood of harmful drinking. Additionally, factors associated with higher socioeconomic status, such as land ownership and residing in pucca houses, were negatively correlated with harmful drinking status. Conversely, higher income and urban residence were positively associated with harmful drinking. The study emphasized the impact of socioeconomic factors and urbanization on alcohol consumption patterns, necessitating targeted interventions for specific socio-economic strata. Recognizing these socio- demographic correlates is pivotal for tailoring intervention and prevention strategies.

In other past research, social peers influence substance abuse, suggesting how social environment may be an effective target for reducing alcohol abuse across a population. A study on alcohol use and abuse among rural adults was conducted in Zimbabwe with the aim of understanding what factors contribute to alcohol abuse in resource-poor countries, particularly investigating the impact of a community popular opinion leader (CPOL) intervention. Results indicated that harmful alcohol consumption correlated with being male, older, unmarried, more educated, of Shona ethnicity, frequent travelers, employed, irreligious, or residing in areas with specific demographic characteristics. Surprisingly, both intervention and control sites experienced significant declines in harmful alcohol use over the study period, with no clear effectiveness of the CPOL intervention observed. Despite this, the study reaffirms the influence of socio-demographic and cultural factors on alcohol use, aligning with patterns seen in other countries (8).

The prevalence of harmful alcohol consumption presents a noteworthy concern especially within the population of PLHIVs, particularly among those who have been on ART for a relatively longer time (9). Disadvantaged and vulnerable populations in SSA face higher rates of alcohol-related deaths and hospitalizations, highlighting the urgency to understand the dynamics of harmful alcohol use especially in the context of HIV care in these communities (10). Therefore, using data from PHIA surveys, we conducted a study to predict harmful alcohol use, and comprehensively understand the relationship between harmful alcohol use and HIV or Antiretroviral Therapy (ART) status while considering the influence of various contributing factors.

## METHODS

### Study design

The initial study design adopted was cross-sectional, followed by subsequent analysis of secondary data extracted from the Population-based HIV Impact Assessment (PHIA) surveys encompassing six Sub-Saharan countries, namely Eswatini, Malawi, Namibia, Tanzania, Zambia, and Zimbabwe.

### Eligibility

We included HIV-positive adults aged 15 to 59 years who participated in the five PHIA surveys, who consented to biomarker testing, who provided complete data on HIV awareness and treatment status and who were classified as receiving ART at the time of the survey either by reporting current ART use or by having detectable blood levels of selected ARVs. We excluded people who reported receiving ART for less than four months to account for people with high baseline VL who may not have had VLS at the time of data collection.

### Data management

R version 4.2 was used for data management. A complete case analysis was adopted, assessment of the nature of missingness was done and analysis was done on complete cases, excluding missing cases. Data management included merging of the data sets, data cleaning and recoding of the data. The primary outcome variable of interest for this study was ‘harmful alcohol use’ a dichotomous variable coded Yes and No. A score of 4 or more will be considered ‘harmful alcohol use’ in males while a score of 3 or more will be considered as ‘harmful alcohol use’ in females. The AUDIT-C used the three standard questions from module 10 of the PHIA questionnaire to generate the scores (11) The explanatory variables included age group, gender (stratified into male and females), marital status (stratified into single, married, widowed and divorced), HIV/ART status (stratified into positive, negative, HIV+ not on ART and HIV- on ART), Area of residence (stratified into rural and urban), highest education level (stratified into none, primary, secondary and tertiary), wealth index (stratified into poorest, poor, rich and richest), relationship to the household head (stratified into head, spouse, child and other), country name (stratified into Eswatini, Malawi, Namibia, Tanzania, Zambia and Zimbabwe).

### Data analysis

Analysis of the data was done in R v4.2. The descriptive data analysis was carried out by calculating frequencies and proportions using stratification as appropriate. The bivariate association between the outcome variable (harmful alcohol use) and predictor variables was assessed using Chi-Square test obtained through univariable logistic regression. The multivariable logistic regression model was fitted using forward and backward model building techniques. In the multivariable model, age group, gender, marital status, HIV/ART status, Area of residence, highest education level, wealth index and relationship to the household head were included in the model based on their P <0.20 at univariable analysis. The logistic regression outputs were reported both as unadjusted (cOR) and adjusted odds ratio (aOR).

We employed to test and apply machine learning (ML) methods through Super Learner, Decision Tree, Random Forest (RF), Lasso Regression, Sample mean and Gradient boosting. Evaluation metrics methods specifically confusion matrix, accuracy, precision, recall, F1 score, and Area under the Receiver Operating Characteristics (AUROC) were used to evaluate the performance of predictive models. The P<0.05 was considered statistically significant.

### Ethical considerations

The initial ethics approval for the study protocols were obtained from the Centers for Disease Control and Prevention Institutional Review Board (IRB), the Columbia University Medical Center IRB, and relevant local regulatory bodies [25] . In our case, we obtained permission to use this data from the International Center for AIDS Care and Treatment Programs (ICAP) at Columbia University. The PHIA datasets were downloaded from https://phia-data.icap.columbia.edu/datasets. As this study used secondary anonymised data, individual informed consent is not required. Furthermore, to conduct this study, we obtained ethics approval from both the University of Zambia Research Ethics committee (UNZABREC Ref. No. 4842-2024) and Zambia National Research Authority (NRA).

## RESULTS

### Respondent characteristics

The characteristics of participants from the six countries are shown in Table 1. The table provides an overview of the study participants’ distribution across the surveyed countries in the PHIA survey, with a total sample size of 14,616 individuals. Notably, Zimbabwe had the largest representation with 3,418 participants, constituting 23.3% of the overall sample. This was followed by Eswatini, with 2,762 participants, and Zambia with 2,268 participants, contributing 18.9% and 15.5% of the total, respectively. Malawi, Namibia, and Tanzania had the least contribution but still substantial proportions, constituting 14.4%, 15.5%, and 12.4% of the overall sample, respectively.

**Table 1:** Sociodemographic characteristics of respondents in Eswatini, Malawi, Namibia, Tanzania, Zambia and Zimbabwe between 2015 and 2019.

|  | Eswatini<br>(N=2762) | Malawi<br>(N=2098) | Namibia<br>(N=2263) | Tanzania<br>(N=1807) | Zambia<br>(N=2268) | Zimbabwe<br>(N=3418) | Overall<br>(N=14616) |
| --- | --- | --- | --- | --- | --- | --- | --- |
| **Age groups** |  |  |  |  |  |  |  |
| 15-19 | 369 (13.4%) | 222 (10.6%) | 286 (12.6%) | 183 (10.1%) | 319 (14.1%) | 381 (11.1%) | 1760 (12.0%) |
| 20-24 | 307 (11.1%) | 222 (10.6%) | 208 (9.2%) | 192 (10.6%) | 271 (11.9%) | 288 (8.4%) | 1488 (10.2%) |
| 25-29 | 343 (12.4%) | 262 (12.5%) | 242 (10.7%) | 192 (10.6%) | 273 (12.0%) | 298 (8.7%) | 1610 (11.0%) |
| 30-34 | 385 (13.9%) | 338 (16.1%) | 262 (11.6%) | 250 (13.8%) | 340 (15.0%) | 477 (14.0%) | 2052 (14.0%) |
| 35-39 | 372 (13.5%) | 336 (16.0%) | 335 (14.8%) | 264 (14.6%) | 327 (14.4%) | 486 (14.2%) | 2120 (14.5%) |
| 40-44 | 248 (9.0%) | 266 (12.7%) | 318 (14.1%) | 212 (11.7%) | 303 (13.4%) | 441 (12.9%) | 1788 (12.2%) |
| 45-49 | 198 (7.2%) | 174 (8.3%) | 236 (10.4%) | 151 (8.4%) | 198 (8.7%) | 318 (9.3%) | 1275 (8.7%) |
| 50-54 | 177 (6.4%) | 125 (6.0%) | 175 (7.7%) | 113 (6.3%) | 142 (6.3%) | 215 (6.3%) | 947 (6.5%) |
| 55+ | 363 (13.1%) | 153 (7.3%) | 201 (8.9%) | 250 (13.8%) | 95 (4.2%) | 514 (15.0%) | 1576 (10.8%) |
| **Gender** |  |  |  |  |  |  |  |
| Male | 1061 (38.4%) | 744 (35.5%) | 809 (35.7%) | 653 (36.1%) | 821 (36.2%) | 1250 (36.6%) | 5338 (36.5%) |
| Female | 1701 (61.6%) | 1354 (64.5%) | 1454 (64.3%) | 1154 (63.9%) | 1447 (63.8%) | 2168 (63.4%) | 9278 (63.5%) |
| **Marital status** |  |  |  |  |  |  |  |
| Single | 1203 (43.6%) | 355 (16.9%) | 1187 (52.5%) | 330 (18.3%) | 602 (26.5%) | 612 (17.9%) | 4289 (29.3%) |
| Married | 1167 (42.3%) | 1217 (58.0%) | 813 (35.9%) | 968 (53.6%) | 1173 (51.7%) | 1956 (57.2%) | 7294 (49.9%) |
| Widowed | 252 (9.1%) | 199 (9.5%) | 106 (4.7%) | 216 (12.0%) | 187 (8.2%) | 516 (15.1%) | 1476 (10.1%) |
| Divorced | 140 (5.1%) | 327 (15.6%) | 157 (6.9%) | 293 (16.2%) | 306 (13.5%) | 334 (9.8%) | 1557 (10.7%) |
| **HIV/ART status** |  |  |  |  |  |  |  |
| Negative | 837 (30.3%) | 503 (24.0%) | 741 (32.7%) | 611 (33.8%) | 696 (30.7%) | 936 (27.4%) | 4324 (29.6%) |
| HIV+ not on ART | 423 (15.3%) | 453 (21.6%) | 212 (9.4%) | 538 (29.8%) | 592 (26.1%) | 711 (20.8%) | 2929 (20.0%) |
| HIV+ On ART | 1502 (54.4%) | 1142 (54.4%) | 1310 (57.9%) | 658 (36.4%) | 980 (43.2%) | 1771 (51.8%) | 7363 (50.4%) |
| **Harmful alcohol use** |  |  |  |  |  |  |  |
| No | 2445 (88.5%) | 1915 (91.3%) | 1673 (73.9%) | 1479 (81.8%) | 1842 (81.2%) | 3106 (90.9%) | 12460 (85.2%) |
| Yes | 317 (11.5%) | 183 (8.7%) | 590 (26.1%) | 328 (18.2%) | 426 (18.8%) | 312 (9.1%) | 2156 (14.8%) |
| **Area of residence** |  |  |  |  |  |  |  |
| Rural | 2117 (76.6%) | 1100 (52.4%) | 1441 (63.7%) | 1084 (60.0%) | 973 (42.9%) | 2378 (69.6%) | 9093 (62.2%) |
| Urban | 645 (23.4%) | 998 (47.6%) | 822 (36.3%) | 723 (40.0%) | 1295 (57.1%) | 1040 (30.4%) | 5523 (37.8%) |
| **Highest Education level** |  |  |  |  |  |  |  |
| None | 211 (7.6%) | 217 (10.3%) | 207 (9.1%) | 370 (20.5%) | 92 (4.1%) | 154 (4.5%) | 1251 (8.6%) |
| Primary | 1033 (37.4%) | 1170 (55.8%) | 831 (36.7%) | 1175 (65.0%) | 889 (39.2%) | 1239 (36.2%) | 6337 (43.4%) |
| Secondary | 886 (32.1%) | 626 (29.8%) | 1175 (51.9%) | 23 (1.3%) | 1107 (48.8%) | 1882 (55.1%) | 5699 (39.0%) |
| Tertiary | 632 (22.9%) | 85 (4.1%) | 50 (2.2%) | 239 (13.2%) | 180 (7.9%) | 143 (4.2%) | 1329 (9.1%) |
| **Wealth index** |  |  |  |  |  |  |  |
| Poorest | 735 (26.6%) | 248 (11.8%) | 794 (35.1%) | 337 (18.6%) | 230 (10.1%) | 871 (25.5%) | 3215 (22.0%) |
| Poor | 625 (22.6%) | 244 (11.6%) | 587 (25.9%) | 357 (19.8%) | 282 (12.4%) | 703 (20.6%) | 2798 (19.1%) |
| Middle | 617 (22.3%) | 294 (14.0%) | 498 (22.0%) | 499 (27.6%) | 488 (21.5%) | 624 (18.3%) | 3020 (20.7%) |
| Rich | 462 (16.7%) | 397 (18.9%) | 293 (12.9%) | 369 (20.4%) | 636 (28.0%) | 632 (18.5%) | 2789 (19.1%) |
| Richest | 323 (11.7%) | 915 (43.6%) | 91 (4.0%) | 245 (13.6%) | 632 (27.9%) | 588 (17.2%) | 2794 (19.1%) |
| **Relationship to the household head** |  |  |  |  |  |  |  |
| Head | 1276 (46.2%) | 1118 (53.3%) | 1007 (44.5%) | 830 (45.9%) | 885 (39.0%) | 1753 (51.3%) | 6869 (47.0%) |
| Spouse | 410 (14.8%) | 511 (24.4%) | 298 (13.2%) | 451 (25.0%) | 615 (27.1%) | 664 (19.4%) | 2949 (20.2%) |
| Child | 537 (19.4%) | 271 (12.9%) | 391 (17.3%) | 262 (14.5%) | 410 (18.1%) | 439 (12.8%) | 2310 (15.8%) |
| Other | 539 (19.5%) | 198 (9.4%) | 567 (25.1%) | 264 (14.6%) | 358 (15.8%) | 562 (16.4%) | 2488 (17.0%) |

**Table 2:** Percentage distribution and correlates of harmful alcohol use in Eswatini, Malawi, Namibia, Tanzania, Zambia and Zimbabwe between 2015 and 2019.

| Variable | No | Yes | Total | P-value |
| --- | --- | --- | --- | --- |
| Gender |  |  |  |  |
| Male | 4,088 (77%) | 1,250 (23%) | 5,338 (100%) | <0.001 |
| Female | 8,372 (90%) | 906 (9.8%) | 9,278 (100%) |  |
| Age group |  |  |  |  |
| 15-19 | 1,679 (95%) | 81 (5%) | 1,760 (100%) | <0.001 |
| 20-24 | 1,307 (88%) | 181 (12%) | 1,488 (100%) |  |
| 25-29 | 1,324 (82%) | 286 (18%) | 1,610 (100%) |  |
| 30-34 | 1,691 (82%) | 361 (18%) | 2,052 (100%) |  |
| 35-39 | 1,746 (82%) | 374 (18%) | 2,120 (100%) |  |
| 40-44 | 1,520 (85%) | 268 (15%) | 1,788 (100%) |  |
| 45-49 | 1,056 (83%) | 219 (17%) | 1,275 (100%) |  |
| 50-54 | 787 (83%) | 160 (17%) | 947 (100%) |  |
| 55+ | 1,350 (86%) | 226 (14%) | 1,576 (100%) |  |
| Area of residence |  |  |  |  |
| Rural | 7,894 (87%) | 1,199 (13%) | 9,093 (100%) | <0.001 |
| Urban | 4,566 (83%) | 957 (17%) | 5,523 (100%) |  |
| HIV/ART status |  |  |  |  |
| Negative | 3,702 (86%) | 622 (14%) | 4,324 (100%) | <0.001 |
| HIV+ not on ART | 2,342 (80%) | 587 (20%) | 2,929 (100%) |  |
| HIV+ On ART | 6,416 (87%) | 947 (13%) | 7,363 (100%) |  |
| Marital status |  |  |  |  |
| Single | 3,678 (86%) | 611 (14%) | 4,289 (100%) | <0.001 |
| Married | 6,213 (85%) | 1,081 (15%) | 7,294 (100%) |  |
| Widowed | 1,307 (89%) | 169 (11%) | 1,476 (100%) |  |
| Divorced | 1,262 (81%) | 295 (19%) | 1,557 (100%) |  |
| Wealth Index |  |  |  |  |
| Poorest | 2,723 (85%) | 492 (15%) | 3,215 (100%) | 0.15 |
| Poor | 2,384 (85%) | 414 (15%) | 2,798 (100%) |  |
| Middle | 2,607 (86%) | 413 (14%) | 3,020 (100%) |  |
| Rich | 2,348 (84%) | 441 (16%) | 2,789 (100%) |  |
| Richest | 2,398 (86%) | 396 (14%) | 2,794 (100%) |  |
| Relationship to the household head |  |  |  |  |
| Head | 5,692 (83%) | 1,177 (17%) | 6,869 (100%) | <0.001 |
| Spouse | 2,613 (89%) | 336 (11%) | 2,949 (100%) |  |
| Child | 2,028 (88%) | 282 (12%) | 2,310 (100%) |  |
| Other | 2,127 (85%) | 361 (15%) | 2,488 (100%) |  |
| Highest education level |  |  |  |  |
| None | 1,044 (83%) | 207 (17%) | 1,251 (100%) | 0.062 |
| Primary | 5,452 (86%) | 885 (14%) | 6,337 (100%) |  |
| Secondary | 4,841 (85%) | 858 (15%) | 5,699 (100%) |  |
| Tertiary | 1,123 (84%) | 206 (16%) | 1,329 (100%) |  |

The prevalence of harmful alcohol use across the entire population was reported at 2,156 individuals (14.8%). Nationally, Namibia stood out with the highest reported harmful alcohol use at 590 individuals (26.1%), while Eswatini, Malawi, and Zimbabwe reported lower rates at 317 individuals (11.5%), 183 individuals (8.7%), and 312 individuals (9.1%), respectively.

Overall age distribution within the surveyed population indicated that the age group 35-39 had the most representation, constituting 1,760 (12.0%) individuals. This pattern was consistent across the six nations under consideration. However, there were variations at country level. In Eswatini, Zambia, and Zimbabwe, the 35-39 age group had the highest representation, constituting 369 (13.4%), 319 (14.1%), and 381 (11.1%) individuals, respectively. On the contrary, the 50-54 age group consistently exhibited the lowest representation, ranging from 6.3% to 6.5%.

Examining gender distribution overall, females constituted the majority across all surveyed countries, representing 9,278 individuals (63.5%) of the overall population. At the country level, Malawi stood out with the highest female representation at 1,354 individuals (64.5%), while Eswatini reported the lowest at 1,701 individuals (61.6%). Males made up the remaining 5,338 individuals (36.5%) overall, with variations ranging from 35.5% in Eswatini to 38.4% in Zimbabwe.

Marital status distribution revealed interesting trends in the surveyed population. The majority fell under the “Married” category, accounting for 7,294 (49.9%) individuals. Zimbabwe had the highest percentage of married individuals at 1,956 individuals (57.2%), while Eswatini and Namibia display significant proportions of “Single” individuals at 1,203 individuals (43.6%) and 1,187 individuals (52.5%), respectively.

A total of 7,363 (50.4%) individuals were identified as HIV positive and actively receiving antiretroviral therapy (ART). At the country level, Zimbabwe and Eswatini demonstrated notably high prevalence among HIV-positive individuals on ART, standing at 1,771 individuals (51.8%) and 1,502 individuals (54.4%), respectively. Malawi, on the contrary, exhibited the highest proportion of HIV-positive individuals not on ART at 453 individuals (21.6%).

The distribution between rural and urban residences was an important aspect to consider. At country level, the majority individuals 9,093 (62.2%) were residing in rural areas. Eswatini and Namibia demonstrated the highest prevalence of rural residence at 2,117 individuals (76.6%) and 1,441 individuals (63.7%), respectively. In contrast, Zambia and Zimbabwe exhibited a higher proportion of urban residents at 1,295 individuals (57.1%) and 1,040 individuals (30.4%), respectively.

Educational attainment is diverse, with the majority having at least a primary education. Nationally, 6,337 individuals (43.4%) have completed primary education, while 5,699 individuals (39.0%) have secondary education. Namibia stands out with the highest proportion having tertiary education at 50 individuals (2.2%), whereas Malawi has the highest representation with primary education at 1,170 individuals (55.8%).

The wealth index categorization provided insights into the socio-economic profiles of the surveyed population. At country level, the “Middle” wealth category was the most prevalent with 3,020 individuals (20.7%). Namibia and Zimbabwe exhibited prominence in this category with 498 individuals (22.0%) and 624 individuals (18.3%), respectively. Zambia, on the other hand, reported the highest representation in the “Rich” and “Richest” categories with 636 individuals (28.0%) and 588 individuals (17.2%), respectively.

Understanding the relationship to the household head was essential. Overall, nearly half of the respondents (47.0%) were identified as the household head, amounted to 6,869 individuals. This trend was consistent across countries, with Eswatini (53.3%) and Zimbabwe (51.3%) reporting the highest percentages of respondents in this category.

### Prevalence of harmful alcohol use

The prevalence of harmful alcohol use across all countries was reported at 14.8%. At country level, Namibia stood out with the highest reported harmful alcohol use with 590 (26.1%) individuals, while Eswatini, Malawi, and Zimbabwe reported lower rates with 317 (11.5%) individuals, 183 (8.7%) individuals, and 312 (9.1%) individuals, respectively.

### Factors associated with harmful alcohol use

#### Bivariate analysis

The chi-square analysis results revealed significant associations between several demographic and health-related variables within the surveyed population. Gender disparities were observed, with a statistically significant differences (p < 0.001). A higher percentage of females (90%) tested positive for HIV compared to males (77%). Similarly, age distribution exhibited statistically significant associations (p < 0.001), with a higher prevalence of HIV-positive individuals found in the older age groups, particularly those aged 55 and above (86%).

Residence in rural or urban areas exhibited significant differences (p < 0.001) concerning HIV status, with a higher percentage of HIV-positive individuals residing in rural areas (87%). Marital status also demonstrated significant associations (p < 0.001), highlighting that married individuals (85%) had a higher prevalence of HIV compared to their single counterparts (86%).

Furthermore, there were substantial differences in HIV prevalence across various relationships to the household head (p < 0.001). Individuals identifying as the household head had a lower HIV prevalence (83%) compared to those in other relationships, such as spouses (89%) or children (88%). Educational attainment did not show significant associations with HIV status, though a trend towards higher prevalence was observed among those with primary education (86%).

Regarding HIV/ART status, statistically significant associations were observed (p < 0.001), indicating that individuals on antiretroviral therapy (ART) had a higher prevalence of HIV (87%) compared to those not on ART (80%). Wealth index categories did not show significant differences in HIV prevalence, suggesting that HIV status was relatively consistent across different wealth strata.

Wealth Index revealed interesting trends despite not showing statistically significant differences in HIV prevalence (p = 0.15). Although not significant, the distribution suggested a potential association between wealth categories and HIV status, with the wealthiest individuals displaying a slightly lower prevalence (86%) compared to the other wealth strata.

Similarly, the variable representing the highest education level did not demonstrate a statistically significant association with HIV prevalence (p = 0.062). However, the data trends suggested a potential inverse relationship, with individuals having no formal education showing a slightly lower HIV prevalence (83%) compared to those with higher educational attainment.

#### A multivariate logistic regression analysis

At the unadjusted level, age demonstrated a significant association with harmful alcohol use. Individuals aged 20-24 had 2.87 times the odds of harmful alcohol use compared to those aged 15-19 (95% CI: 2.20- 3.78, p < 0.001). The odds increased across subsequent age groups, reaching 4.44 for individuals aged 35- 39 (95% CI: 3.48-5.73, p < 0.001). These associations remained significant in the adjusted analysis, where individuals aged 20-24 exhibited 3.97 times the odds of harmful alcohol use (95% CI: 3.01-5.27, p < 0.001), and the odds continued to rise with age, with individuals aged 55 and above having 6.77 times the odds (95% CI: 5.06-9.14, p < 0.001).

Gender differences were evident, with females showing a protective effect in both unadjusted (Odds Ratio [OR] = 0.35, 95% CI: 0.32-0.39, p < 0.001) and adjusted analyses (Adjusted Odds Ratio [AOR] = 0.32, 95% CI: 0.29-0.35, p < 0.001). Area of residence played a role, where individuals in urban areas had 1.38 times the odds of harmful alcohol use compared to those in rural areas at the unadjusted level (95% CI: 1.26- 1.51, p < 0.001). This association strengthened in the adjusted analysis, with individuals in urban areas now having 1.74 times the odds (95% CI: 1.54-1.98, p < 0.001).

HIV/ART status showed association. At the unadjusted level, being HIV-positive but not on ART increased the odds of harmful alcohol use (OR = 1.49, 95% CI: 1.32-1.69, p < 0.001), while being HIV-positive on ART decreased the odds (OR = 0.88, 95% CI: 0.79-0.98, p = 0.02). In the adjusted analysis, the association with being HIV-positive on ART remained significant, with an AOR of 0.65 (95% CI: 0.57-0.73, p < 0.001). Wealth index dynamics were also influential. At the unadjusted level, individuals in the middle wealth index category had 28% lower odds of harmful alcohol use (OR = 0.72, 95% CI: 0.62-0.83, p < 0.001), and those in the richest category had 42% lower odds (OR = 0.58, 95% CI: 0.48-0.69, p < 0.001) compared to the poorest category. These associations persisted in the adjusted analysis.

Marital status demonstrated varied associations. Being married showed a protective effect in both unadjusted (OR = 1.05, 95% CI: 0.94-1.17, p = 0.4) and adjusted analyses (AOR = 0.6, 95% CI: 0.52-0.68, p < 0.001). Widowed individuals exhibited lower odds of harmful alcohol use at the unadjusted level (OR = 0.78, 95% CI: 0.65-0.93, p = 0.007) and the adjusted level (AOR = 0.7, 95% CI: 0.56-0.86, p < 0.001).

Divorced individuals were significantly associated with harmful alcohol use at the unadjusted level (OR = 1.41, 95% CI: 1.21-1.64, p < 0.001), but this association became non-significant in the adjusted analysis (AOR = 0.96, 95% CI: 0.81-1.15, p = 0.7).

### Predictive modelling of harmful alcohol use

We fitted the logistic regression on the training dataset which contained 70% of the dataset. The final model of the logistic regression (see Table 3) was used to determine the confusion matrix for the training dataset. The output of the logistic regression analysis using the test dataset is shown in Figure 3. Based on the predicted logistic regression model, the key determinants of the harmful alcohol use were: gender, area of residence, HIV/ART status, marital status, and country of residence. After training the data, we used Super Learner, Decision Tree, Random Forest (RF), Lasso Regression, Sample mean and Gradient boosting on the test data and the model performance results are shown in Figure 3. Based on these models, the best performing models were Lasso or Super Learner or Random Forest were the best performing models while gradient boosting models or sample mean did not perform well.

**Figure 1:**
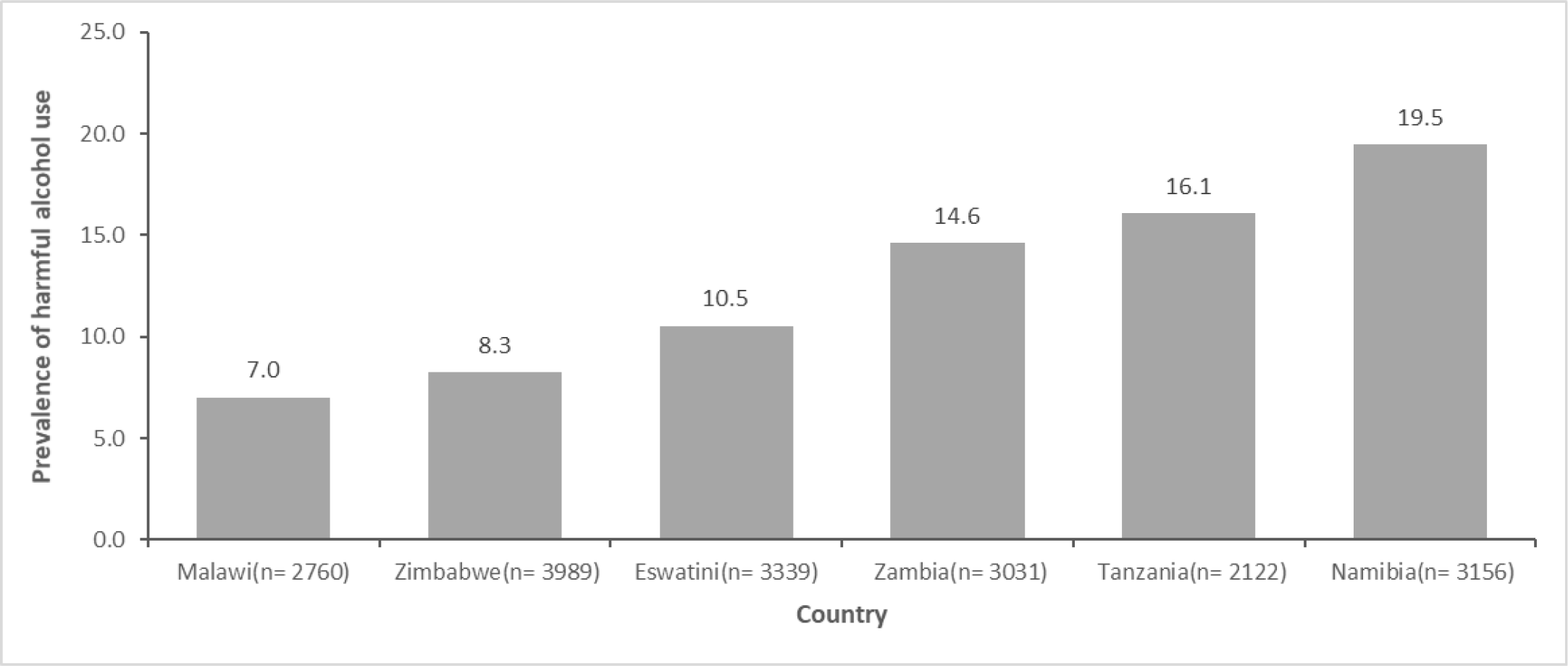
Prevalence of harmful alcohol use in Eswatini, Malawi, Namibia, Tanzania, Zambia and Zimbabwe between 2015 and 2017

**Figure 2:**
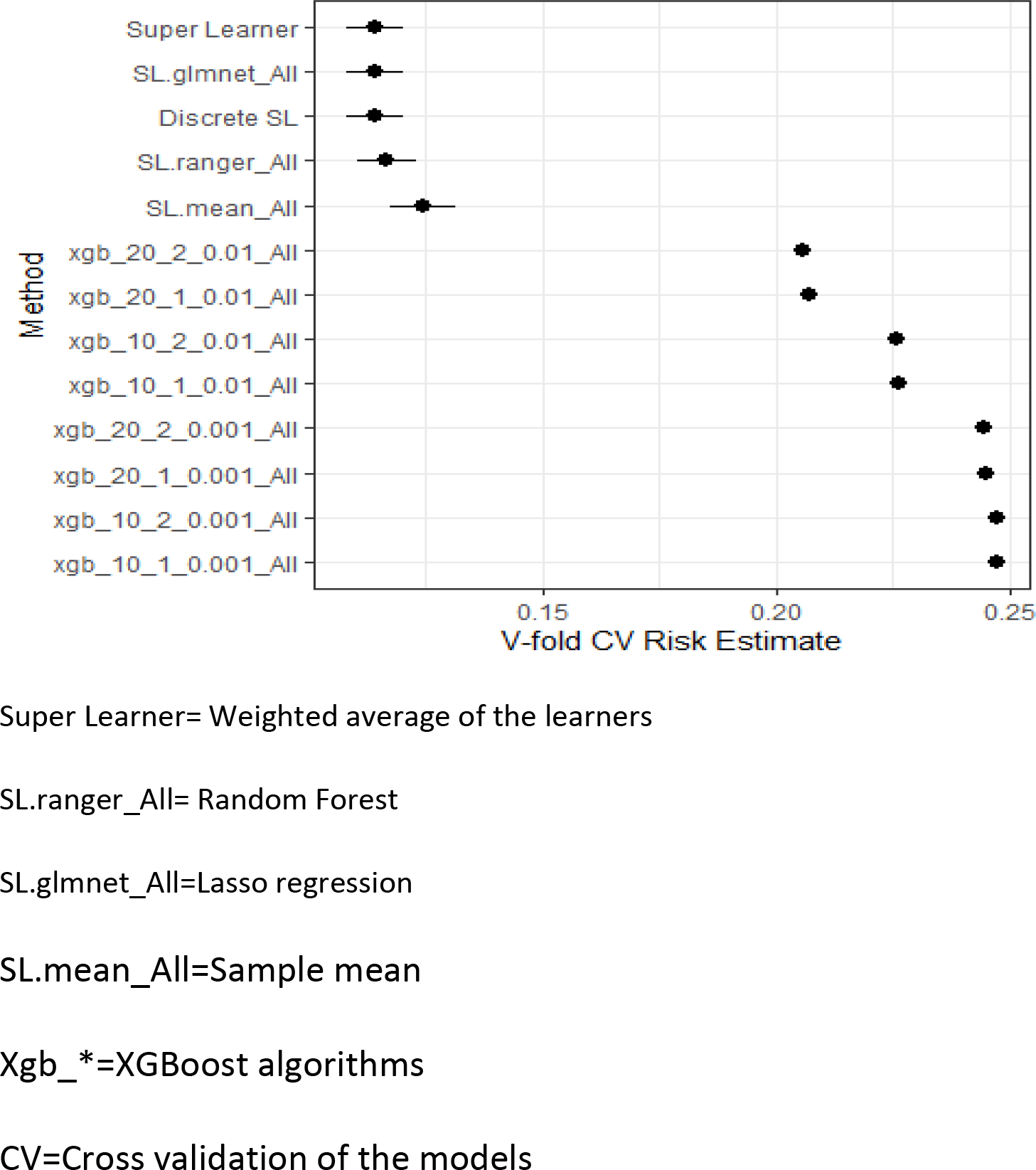
Plot the performance for the different supervised machine learning models fitted

**Figure 3:**
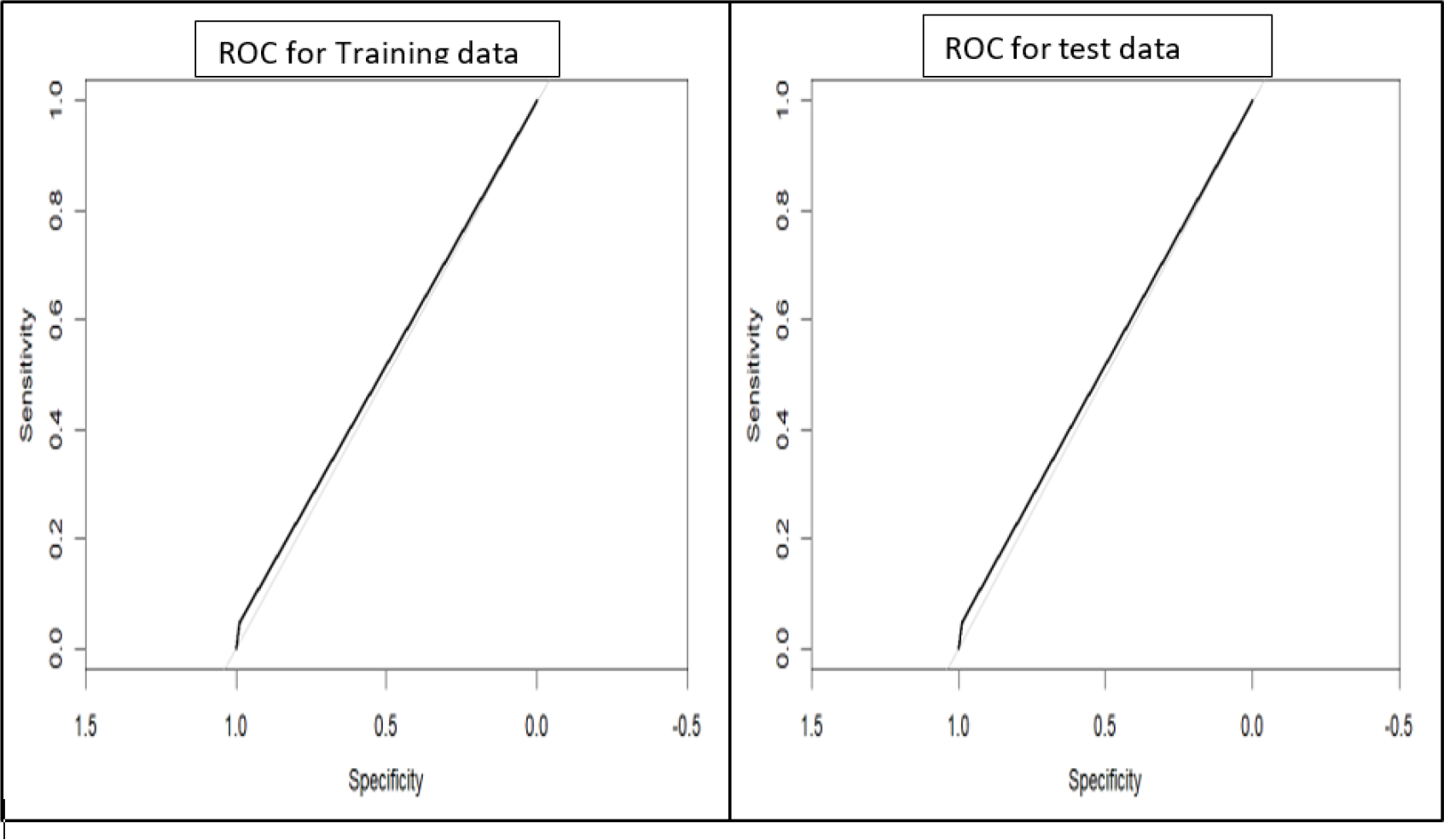
R**O**C **curves for training and testing curves for screening harmful alcohol use in Eswatini, Malawi, Namibia, Tanzania, Zambia and Zimbabwe between 2015 and 2017**

**Table 3:**
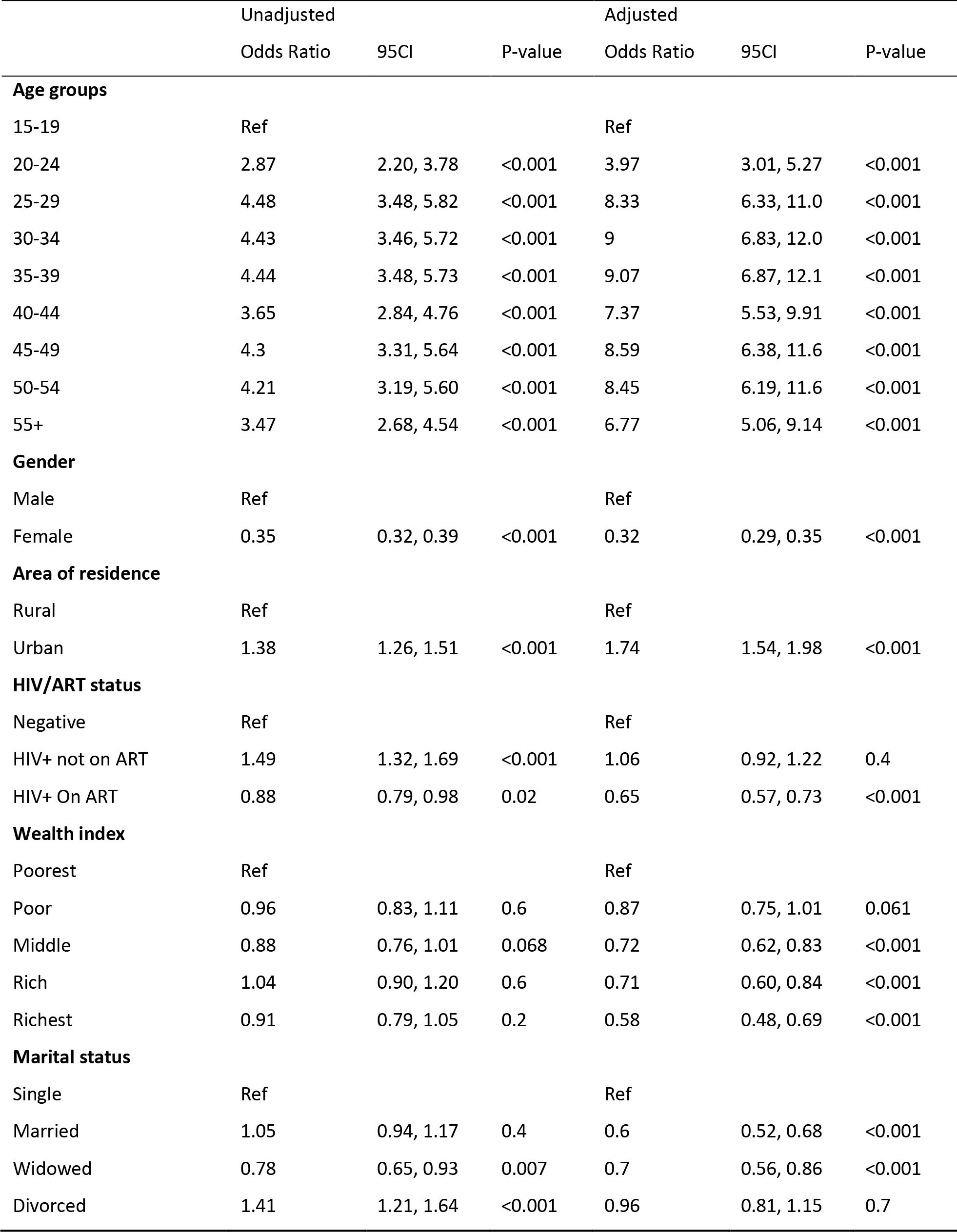
Univariable and multivariable logistic regression analysis of harmful alcohol use in in Eswatini, Malawi, Namibia, Tanzania, Zambia and Zimbabwe between 2015 and 2019.

### Assessment of the lasso model

The metrics for assessing the lasso regression model are shown in Figure 3, Table 4 and Table 5. In the presented ROC analysis, the model exhibited consistently high accuracy in the test and training datasets was 0.85. The precision values, representing the accuracy of positive predictions, were 0.52 for the test dataset.

**Table 4:** Confusion matrix for the accuracy of the fitted model of choice in classifying individuals to the harmful alcohol use in Eswatini, Malawi, Namibia, Tanzania, Zambia and Zimbabwe between 2015 and 2019.

| Test data |  |  |  |
| --- | --- | --- | --- |
| Harmful alcohol consumption | No | Yes | Total |
| No | 3691 | 47 | 3738 |
| Yes | 619 | 28 | 647 |
| Total | 4310 | 75 | 4385 |

**Table 5:** Performance metrics for predicting harmful alcohol use in Eswatini, Malawi, Namibia, Tanzania, Zambia and Zimbabwe between 2015 and 2017.

| Parameter | Value |
| --- | --- |
| Precision | 0.52 |
| Accuracy | 0.85 |
| Recall | 0.01 |
| False Positive Rate | 0.01 |
| False Negative Rate | 0.95 |
| F1 Score | 0.02 |

### Predicted multiple logistic regression parameters

Figure 4 shows the predictors of harmful alcohol use based on test dataset for the Lasso Regression. The output was consistent with the conventional data analysis shown in Table 3. The odds were found to increase with age, with those between the ages of 30 and 34 showing the highest risk. The predictions for females’ odds were substantially lower than those for males, suggesting a protective effect, and these differences were significant (P<0.0001). Living in an urban area was linked to an increased risk of harmful alcohol use, with those who live there expected to have a higher chance of consuming alcohol in a harmful way than individuals who live in rural areas. Those who were HIV-positive and not receiving ART were at a higher risk of engaging in harmful alcohol use. Additionally, divorced individuals had a higher of alcohol consuming alcohol compared to those in marriage and the single.

**Figure 4:**
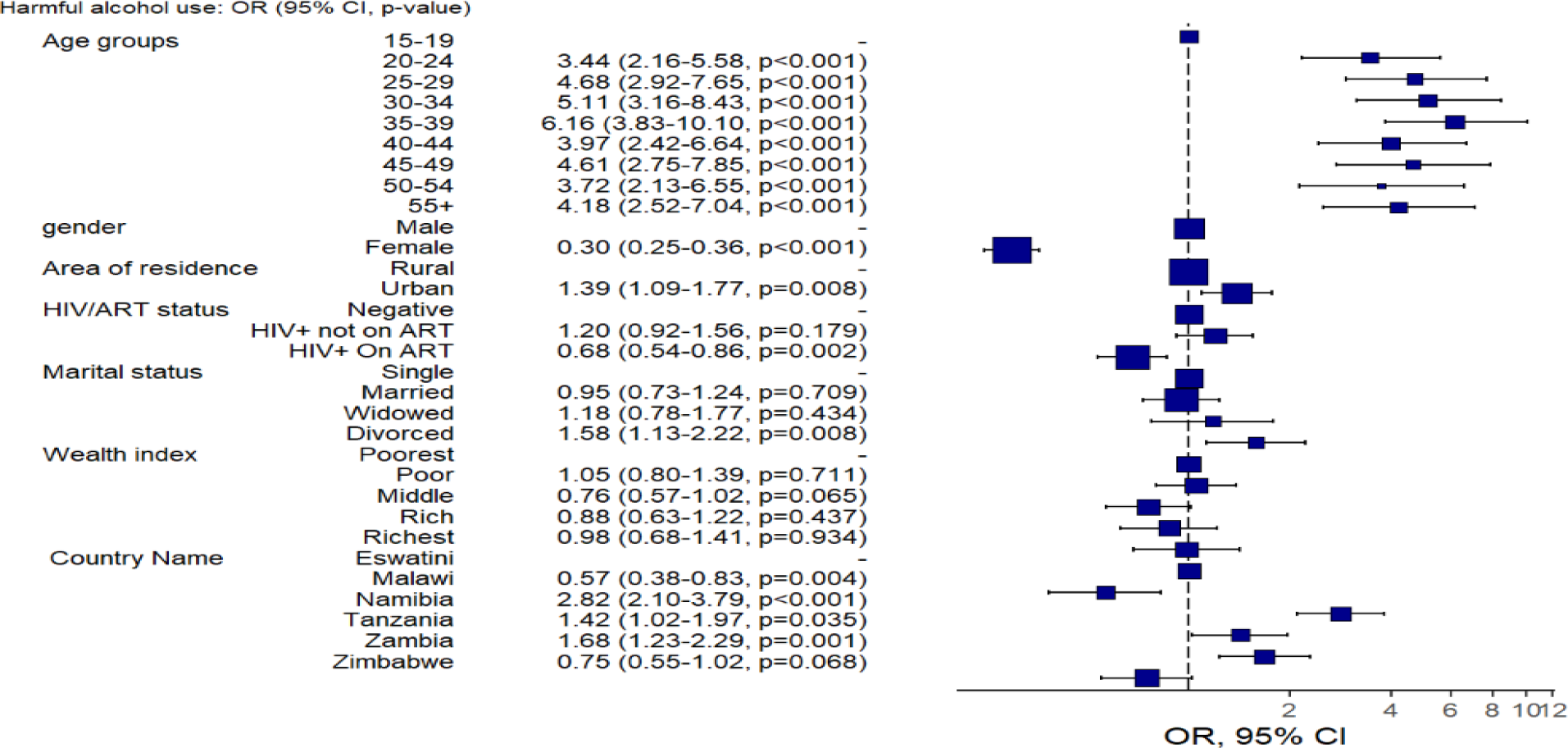
Predictive model of the outcomes using logistic regression and the forest plot of odds ratio of harmful alcohol use in Eswatini, Malawi, Namibia, Tanzania, Zambia and Zimbabwe between 2015 and 2019

## DISCUSSION

The study provided valuable insights into the prevalence of harmful alcohol consumption in six countries from 2015 to 2019. Namibia had the highest prevalence rate at 18.5%, representing a significant public health problem. In contrast, Ethiopia had a lower prevalence rate of 7.2%, suggesting possible cultural or contextual factors influencing alcohol consumption behavior. This finding resonates with existing literature highlighting elevated alcohol consumption rates in certain sub-Saharan African regions, emphasizing the need for targeted interventions in Namibia to address this pressing issue (12).

Consistent with earlier studies, the demographic analysis revealed specific patterns associated harmful alcohol consumption. Studies on alcohol consumption have consistently shown that age increases the odds of alcohol consumption. Younger age groups, particularly those between the ages of 20-24 showed lower odds, highlighting how vulnerable older age groups are. This is consistent with the trends identified in the literature, where risky drinking tends to peak in middle age (13).

The study’s findings regarding gender disparities validate the widespread belief that men are more likely than women to use harmful alcohol (5). Gender-sensitive interventions are crucial, as evidenced by the protective effect that females display, with their odds reduced by 12.3%. Contrary to literature highlighting a negative association between education and alcohol use, this study did not find significant associations. This deviation may be attributed to cultural differences and warrants further investigation.

Interesting results were obtained when the analysis was adjusted for HIV/ART status. Individuals on Antiretroviral Therapy (ART) displayed a decreased risk of harmful alcohol use, with odds reduced by 30.2%. This aligns with studies suggesting a complex interplay between HIV management and substance use (14). The protective effect of ART emphasizes the potential dual benefits of HIV treatment programs in addressing both infectious and behavioral health challenges.

Socioeconomic factors, represented by the wealth index, demonstrated noteworthy associations. Middle-income individuals exhibited lower odds of harmful alcohol use (14.8% lower odds), indicating potential disparities across socioeconomic strata. This aligns with research showing how socioeconomic status influences alcohol consumption patterns (15). Variations by country, especially Namibia’s higher odds, highlight how crucial it is to customize interventions to unique sociocultural contexts.

Chi-square analyses revealed a strong correlation between the use of harmful alcohol and living in an urban area, with those who live in urban areas having higher odds (23.6% higher odds). This discrepancy between urban and rural areas is consistent with the association between urbanization and alcohol consumption reported in several studies (6). The influence of marital status was also significant; married individuals demonstrated a protective effect (17.1% lower odds), while divorced individuals showed increased odds (29.8% higher odds). These findings underscore the need for targeted interventions addressing the unique vulnerabilities of urban populations and divorced individuals.

The logistic regression outputs further explained the distinct relationships between demographic variables and harmful alcohol use. Age remained a strong predictor, with increasing odds observed in older age groups. Gender disparities persisted, with males exhibiting higher odds (19.5% higher odds), reinforcing the importance of gender-sensitive interventions. Despite the lack of significant associations in descriptive analyses, logistic regression highlighted the complexity of the relationship between education and harmful alcohol use, calling for further exploration.

Machine learning models provided valuable predictive insights into harmful alcohol use. The established logistic regression equation used in the model demonstrated high accuracy in identifying at-risk individuals. This analysis underscored the significance of age, gender, and HIV/ART status in predicting harmful alcohol use, aligning with the findings from traditional statistical analyses.

## Conclusion

This study used a multi-country approach to investigate harmful alcohol use in Sub-Saharan African countries using PHIA surveys conducted between 2015 and 2019. The research aimed to identify the factors associated with harmful alcohol use. With significant differences between the six countries under study, the prevalence findings highlighted the serious public health threat that excessive alcohol consumption poses. Particularly for Namibia, the alarming prevalence rate of 18.5% highlights the need for focused interventions. Ethiopia, on the other hand, showed a prevalence rate that was lower at 7.2%, suggesting that cultural or contextual factors may have an impact on patterns of alcohol consumption. The complex relationships between age, gender, marital status, and education with harmful alcohol use were examined through demographic analyses. The need for age tailored interventions is reinforced by the age-specific analysis, where older age groups showed higher odds. The need for gender-sensitive approaches is highlighted by gender disparities, where men showed higher odds. Unexpectedly, the study did not find a substantial link between drinking alcohol negatively and education, indicating the need for more research. The analysis gained a new perspective with the inclusion of health-related variables, specifically HIV/ART status.

## Strengths and limitations

The study had a strong multi-country representation, using six different countries with varying sociocultural contexts. This inclusiveness improved the findings’ generalizability. The study took a comprehensive approach by incorporating demographic variables with health-related factors like HIV/ART status. This enhanced understanding of the causes and effects of harmful alcohol consumption. This study was noteworthy for its innovative use of machine learning model. The study’s conclusions are useful outside of the academic setting because they provide policymakers and public health professionals with doable suggestions. The study’s reference period, covering data collected between 2015 and 2019, might not capture more recent developments in harmful alcohol consumption patterns.

## AUTHORS’ CONTRIBUTIONS

MG led the manuscript writing, WFN conducted data management and analysis; CZ advised on the data analysis and policy insights on the paper. All authors read and approved the final manuscript.

## Data Availability

The data used in this manuscript are derived from the Population-based HIV Impact Assessment (PHIA). Please refer to the data access policies of PHIA for more information

## ACKNOWLEDGEMENT

The authors would like to thank the ICAP at Columbia University for allowing us to use the data.

